# NIH Funding of COVID-19 Research in 2020: a Cross Sectional Study

**DOI:** 10.1101/2021.12.08.21267482

**Authors:** Logesvar Balaguru, Chen Dun, Andrea Meyer, Sanuri Hennayake, Christi M. Walsh, Christopher Kung, Brittany Cary, Frank Migliarese, Tinglong Dai, Ge Bai, Kathleen Sutcliffe, Martin A. Makary

**Affiliations:** Department of Surgery, School of Medicine, Johns Hopkins University, Baltimore, MD; Pennsylvania State College of Medicine, Hershey, PA; MedStar Washington Hospital Center, Washington, DC; Carey Business School, Johns Hopkins University, Baltimore, MD; Bloomberg School of Public Health, Johns Hopkins University, Baltimore, MD; Department of Anesthesia and Critical Care Medicine, School of Medicine, Johns Hopkins University, Baltimore, MD; School of Nursing, Johns Hopkins University, Baltimore, MD

## Abstract

**Objective:** This study aims to characterize and evaluate the NIH’s grant allocation pattern of COVID-19 research.

**Design:** Cross sectional study

**Setting:** COVID-19 NIH RePORTER Dataset was used to identify COVID-19 relevant grants.

**Participants:** 1,108 grants allocated to COVID-19 research.

**Main Outcomes and Measures:** The primary outcome was to determine the number of grants and funding amount the NIH allocated for COVID-19 by research type and clinical/scientific area. The secondary outcome was to calculate the time from the funding opportunity announcement to the award notice date.

**Results:** The NIH awarded a total of 56,169 grants in 2020, of which 2.0% (n=1,108) were allocated for COVID-19 research. The NIH had a $42 billion budget that year, of which 5.3% ($2.2 billion) was allocated to COVID-19 research. The most common clinical/scientific areas were social determinants of health (n=278, 8.5% of COVID-19 funding), immunology (n=211, 25.8%), and pharmaceutical interventions research (n=208, 47.6%). There were 104 grants studying COVID-19 non-pharmaceutical interventions, of which 2 grants studied the efficacy of face masks and 6 studied the efficacy of social distancing. Of the 83 COVID-19 funded grants on transmission, 5 were awarded to study airborne transmission of COVID-19, and 2 grants on transmission of COVID-19 in schools. The average time from the funding opportunity announcement to the award notice date was 151 days (SD: ±57.9).

**Conclusion:** In the first year of the pandemic, the NIH diverted a small fraction of its budget to COVID-19 research. Future health emergencies will require research funding to pivot in a timely fashion and funding levels to be proportional to the anticipated burden of disease in the population.

## Introduction

The National Institutes of Health (NIH) is the world’s largest funder of biomedical research, employing over 20,000 people with a $41.7 billion budget in 2020 appropriated by Congress.^1,2^ NIH-sponsored research aims to tackle the toughest problems in healthcare while financially supporting research at every stage. Prior research suggested that the NIH research funding has been disproportionately aligned to disease burden in the population.^3–6^ Throughout the 1990s, NIH funding patterns were under major scrutiny from Congress and the scientific community due to concerns that funding allocations by the NIH failed to adequately reflect the burden of disease on society.^5^ In 1998, the (IOM) released a groundbreaking report guiding the NIH to improve and develop disease-specific funding processes.^20^ A landmark study published in the *New England Journal of Medicine* as well as a follow-up study by Gillum et. al in 2011 revealed that the NIH disease-specific funding levels were not correlated with several measures of disease burden.^4,5^

The COVID-19 pandemic tested the NIH’s ability to fund critical research to answer research questions that significantly affect public health and require urgent scientific clarity. We analyzed the relative weight and composition of the NIH research funding of COVID-19 research in 2020 to evaluate the responsiveness of the agency to the pandemic.

## Methods

### Study Design and Settings

We conducted a cross sectional study using the NIH Research Portfolio Online Reporting Tools Expenditures and Results (RePORTER) datasets of all COVID-19 grants, including grants created by a special COVID-19 appropriations by congress.^7,8^ We also reviewed the NIH Fiscal Year 2020 budget to identify spending on NIH COVID-19 research.^9^

We reviewed all grants funded for COVID-19 research between January 1, 2020, and December 31, 2020. For each grant, we collected the date of funding opportunity announcement, award notice date, and the amount awarded as listed in the NIH RePORTER dataset. The date of the Funding Opportunity Announcement was obtained from the NIH COVID-19 grant opportunities.^10^

We categorized each grant into one of six research types: basic science, clinical science, translational science, public health, infrastructure & education, and other (Appendix 1). Each NIH-funded grant was also subcategorized by clinical/scientific area (Appendix 2). We adapted definitions for research areas and subcategories of primary research subjects from NIH Research, Condition, and Disease Categorization (RCDC) thesaurus and supplemented them using definitions from the Association of American Medical Colleges (AAMC), National Cancer Institute (NCI), Economic Social Research Council, the Department of Health and Human Services (DHHS), and *Methods in Educational Research*.^11–17^

Each grant was independently reviewed and categorized by at least two independent reviewers (LB, SH, CD, CK, AM, BC). For grants that were categorized differently, a study group discussed the aims of the grant and made a final decision.

### Data Source

RePORTER is an electronic tool developed by the NIH that works in conjunction with the NIH’s RePORT website. This tool allows users to generate lists of funded NIH studies based on specific search criteria, such as funding source and research area.^9^ To obtain a list of all the grants that funded COVID-19 research in 2020, we used the NIH’s pre-generated COVID-19 RePORTER dataset.^7^ The information describing 2020 NIH funding by research was found on the Research, Condition, and Disease Categories (RCDC) RePORTER database. ^18^

### Outcomes

The primary outcome for this analysis was to calculate the number of grants and funding the NIH allocated toward COVID-19 in 2020 to the 6 research types and each clinical/scientific area. The secondary outcome was to calculate the time from funding opportunity announcement to award.

### Statistical Analysis

We calculated the funding amount for research areas by compiling each grant’s total funding amount allocated by the NIH. The funding amount for the clinical/scientific area was calculated based on each grant’s categorization. We plotted the weekly number of COVID-19 grants awarded during 2020. Data cleaning and statistical analyses were conducted using Stata (Version 14.0).

## Results

In 2020, COVID-19 research accounted for 5.3% ($2.2 billion) of the annual NIH budget of $41.7 billion.^19^ Of the $2.2 billion that the NIH spent on COVID-19 research, 86.5% was allocated from congressional special appropriations while the remaining 13.5% of COVID-19 funding originated from the regular NIH annual budget that year. We found that several disease and condition-specific research areas were funded at levels much greater than COVID-19 (Figure 1). Rare Diseases research received 2.5-fold more funding than coronavirus research and aging research received 2.2-fold more research funding than coronavirus research.^18^

**Figure 1:**
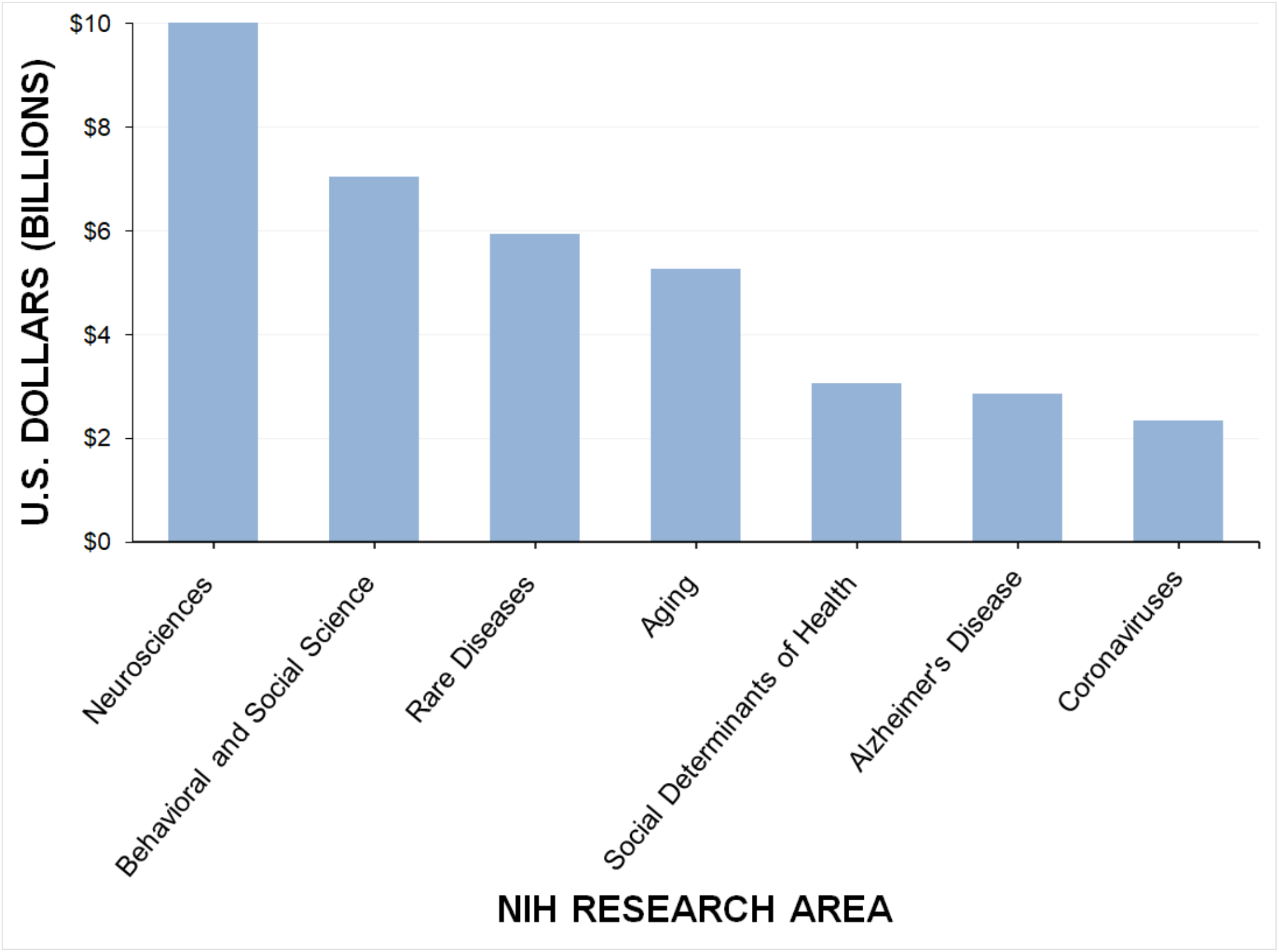
NIH Funding by Research Area (2020)

There were 1,419 NIH COVID-19 grants from the year 2020 in the NIH RePORTER dataset. Of these, we identified 1,108 COVID-19 grants with relevance to COVID-19 research, 24 were duplicates appearing in different places and 287 did not have COVID-19 relevance. Of the 1,108 COVID-19 grants identified, 266 grants were able to be matched to their funding opportunity announcement. The average COVID-19 grant was issued funding 151 days (SD: ±57.9) after its funding opportunity announcement, with a median of 137 days (IQR: 109-196) and range from 43-295 days.

In the first three months of the global pandemic, a total of 6 grants were awarded for COVID-19 research. In the first half of 2020, a total of 240 grants were awarded funding (Figure 2). Accordingly, in the first three months of 2020, the NIH spent a total of 0.1% of its annual budget on COVID-19 research. In the first half of 2020, the NIH spent 1.2% of its annual budget on COVID-19 research. The months with the most COVID-19 research grants awarded were August and October.

**Figure 2.**
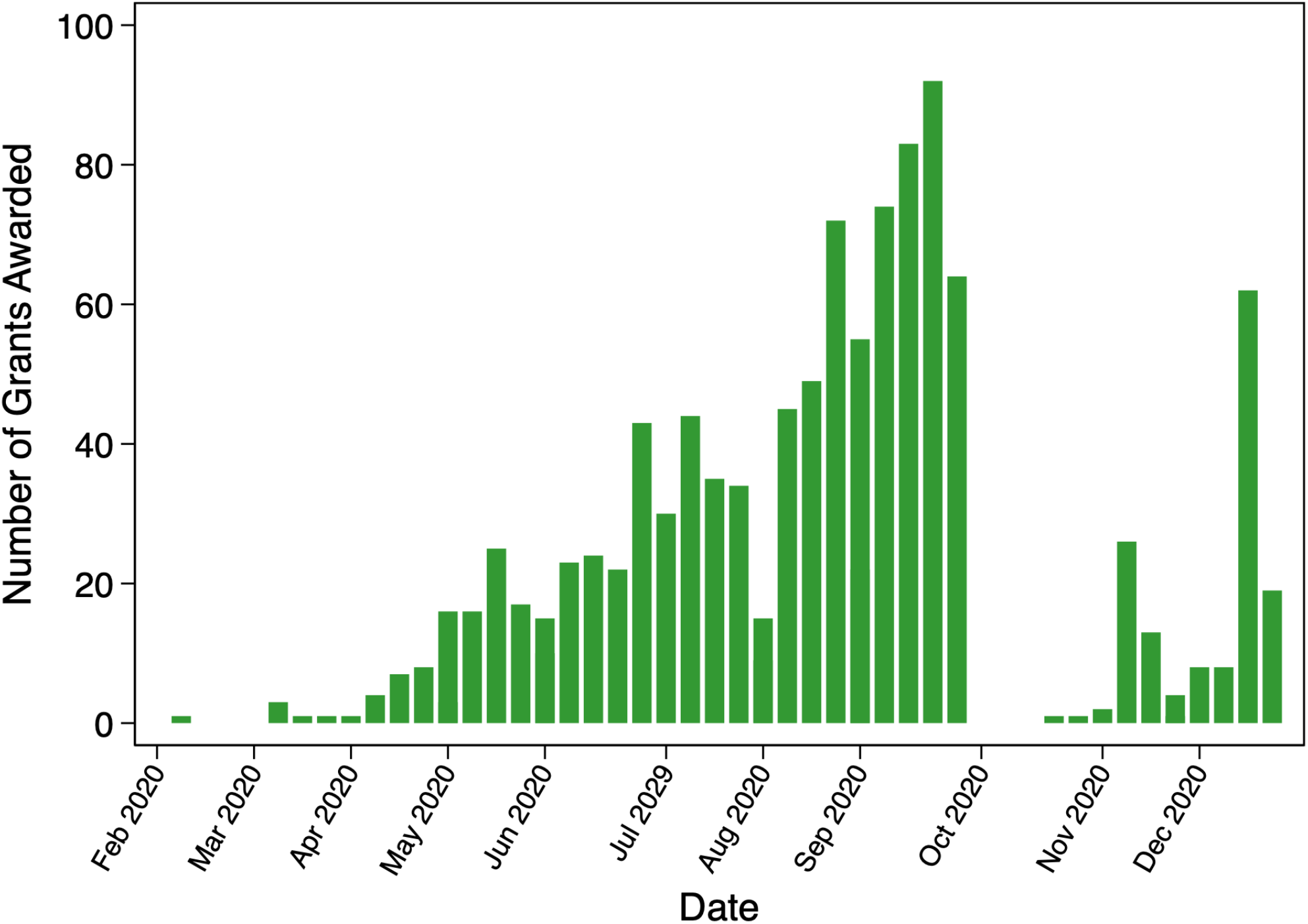
Number of COVID-19 Grants Approved by NIH in 2020.

Regarding the type of the COVID-19 research funded, basic science research comprised the greatest number of grants funded by the NIH with a total of 313 grants, compromising 6.9% of total COVID-19 research funding. There were 231 grants awarded for public health research and 231 grants awarded for clinical research, accounting for 5.7% and 26.8% of NIH COVID-19 funding, respectively. The NIH allocated the largest dollar amount to infrastructure and education research with 55.5% of all COVID-19 funds going to these purposes with 216 grants, accounting for 3.0% of the NIH’s annual budget (Table 1).

**Table 1:** NIH Grants for COVID-19 Research by Research Type (2020)

|  | Number of<br>COVID-19<br>Grants (%) | Dollars Spent,<br>USD | Percent of All<br>COVID-19<br>Funding (%) | Percent of Total<br>NIH Annual<br>Budget (%) |
| --- | --- | --- | --- | --- |
| Basic Science | 313 (28.25%) | 151,252,564 | 6.85 | 0.36 |
| Translational | 81 (7.31%) | 85,436,684 | 3.87 | 0.20 |
| Clinical | 231 (20.85%) | 591,533,574 | 26.77 | 1.42 |
| Infrastructure and Education | 216 (19.49%) | 1,235,403,053 | 55.92 | 2.96 |
| Public Health | 231 (20.85%) | 124,813,879 | 5.65 | 0.30 |
| Other | 36 (3.25%) | 20,946,874 | 0.95 | 0.05 |
| Total | 1,108 | 2,209,386,628 | 100.00 | 5.30 |

The most common clinical/scientific areas of research were social determinants of health (n=278 grants, 8.5% of COVID-19 funding), immunology (n=211 grants, 25.8% of COVID-19 funding), and pharmaceutical interventions (n=208 grants, 47.6% of COVID-19 funding) (Table 2). Of the 208 grants dedicated to pharmaceutical intervention research, 85 grants focused on novel therapeutics development (6.4% of COVID-19 funding), 79 grants focused on existing therapeutics (28.2% of COVID-19 funding), and 69 grants on vaccine development (32.2% of COVID-19 funding). Of the 211 immunology grants, 41 grants studied immunity gained after infection of COVID-19, and 15 grants studied immune response from vaccination. Of 64 neurological grants, 13 grants focused on changes of tastes or smell.

**Table 2:**
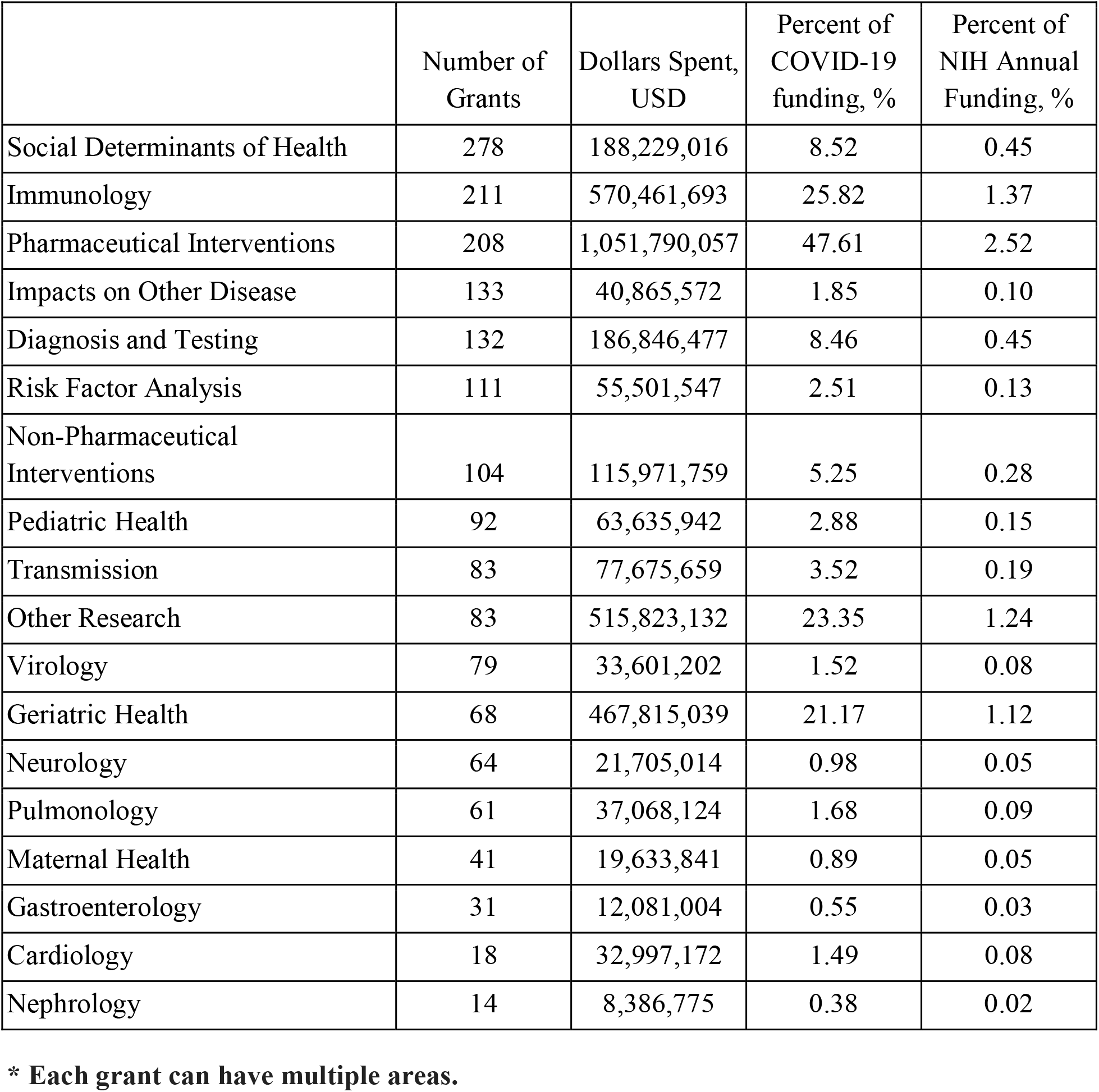
NIH Grants for COVID-19 by Clinical/Scientific Area (2020)*

|  | Number of Grants | Dollars Spent, USD | Percent of COVID-19 funding, % | Percent of NIH Annual Funding, % |
| --- | --- | --- | --- | --- |
| Social Determinants of Health | 278 | 188,229,016 | 8.52 | 0.45 |
| Immunology | 211 | 570,461,693 | 25.82 | 1.37 |
| Pharmaceutical Interventions | 208 | 1,051,790,057 | 47.61 | 2.52 |
| Impacts on Other Disease | 133 | 40,865,572 | 1.85 | 0.10 |
| Diagnosis and Testing | 132 | 186,846,477 | 8.46 | 0.45 |
| Risk Factor Analysis | 111 | 55,501,547 | 2.51 | 0.13 |
| Non-Pharmaceutical Interventions | 104 | 115,971,759 | 5.25 | 0.28 |
| Pediatric Health | 92 | 63,635,942 | 2.88 | 0.15 |
| Transmission | 83 | 77,675,659 | 3.52 | 0.19 |
| Other Research | 83 | 515,823,132 | 23.35 | 1.24 |
| Virology | 79 | 33,601,202 | 1.52 | 0.08 |
| Geriatric Health | 68 | 467,815,039 | 21.17 | 1.12 |
| Neurology | 64 | 21,705,014 | 0.98 | 0.05 |
| Pulmonology | 61 | 37,068,124 | 1.68 | 0.09 |
| Maternal Health | 41 | 19,633,841 | 0.89 | 0.05 |
| Gastroenterology | 31 | 12,081,004 | 0.55 | 0.03 |
| Cardiology | 18 | 32,997,172 | 1.49 | 0.08 |
| Nephrology | 14 | 8,386,775 | 0.38 | 0.02 |

**\* Each grant can have multiple areas.**

There were 132 grants awarded for COVID-19 testing, compromising 8.5% of all COVID-19 funding. There were 83 grants on COVID-19 transmission, representing 3.5% of COVID-19 funding. Of these, 5 studied airborne transmission and 2 grants studied COVID-19 transmission in schools.

A total of 104 grants focused on non-pharmaceutical interventions, with 6 grants on the efficacy of social distancing and 2 grants on the efficacy of face masks. Additionally, 92 grants studied the effects of COVID-19 infection in pediatric populations 10 of which examined Inflammatory Syndrome in Children (MIS-C). Geriatric health and COVID-19 was awarded 68 grants and maternal health and COVID-19 was awarded 41 grants. There were no grants dedicated to studying the efficacy of face masks in children.

## Discussion

Despite the escalating public health threat and poorly-understood mechanism of transmission of the novel coronavirus in 2020, the NIH only spent 5.3% of their total budget that year on COVID-19 research, extending prior literature that that NIH funding priorities misaligned with disease burden in the population.^5,20^ The NIH’s slow start in funding COVID-19 research was also noted in a February 2021 study in *Health Affairs* by Sampat and Shadlen.^6^ They described the current low investment in COVID-19 research as “small compared with the potential value of these interventions for ameliorating or preventing the disease and securing a return to normalcy”. A stronger research effort could have helped reduce transmission of the infection before a vaccine became available.

Infrastructure and education accounted for 55.9% of NIH COVID-19 funding, yet many of the major clinical questions surrounding COVID-19 transmission were unanswered at that time, such as transmission among children. Significant restrictions have been placed on the nation’s 52 million school-aged children, including school closures, 6-foot distancing requirements, and outdoor masking while distancing; however, only a few grants were dedicated to study these questions in this unique population, creating challenges for evidence-based policymaking.

The lack of rapid clinical research funding to understand COVID-19 transmission may have contributed to the politicization of the virus. Some of the most basic questions that were being asked of medical professionals in early 2020, such as how it spreads, when infected individuals are most contagious, and whether masks protect individuals from spreading or getting the virus, went unanswered. In the absence of evidence-based answers to the common questions the public was asking, political opinions filled that vacuum.

The social and political climate of the COVID-19 pandemic has been plagued with misinformation hindering important mitigation efforts. The NIH, as the largest research funding arm of the federal government, has a responsibility to fund research that can address misinformation with evidence. A resilient health care system in times of crisis should be able to pivot funding toward specific grants answering critical gaps in knowledge. NIH may consider developing procedures to rapidly pivot funding and guidelines for reviewing targeted proposals relevant to addressing a public health emergency.

Our study has several limitations. The type of research and the clinical/scientific areas studied were based on definitions that may not be collectively exhaustive and mutually exclusive. In addition, we only reviewed abstracts and did not review the entire funded proposals. There were other barriers to clinical research that were not captured here, including slow institutional review boards and long journal peer-review times. A rapid research protocol that protects research subjects with standard ethical principles for research could be developed for the next health emergency.

## Conclusion

NIH funding patterns for COVID-19 grants did not align with COVID-19 disease burden and were allocated slowly. The NIH should develop mechanisms to rapidly pivot funding to address scientific unknowns associated with a sudden, large-scale health emergency. Supporting sound clinical research aimed at developing evidence-based recommendations is important for public policy and promotes public trust in the medical profession during a pandemic.

### Strengths and limitations of this study

- Our study characterized and evaluated the NIH’s grant allocation pattern of COVID-19 research in the year of 2020.
- We categorized 1,108 grants by research type and clinical/scientific area and identified NIH funding gaps in research dedicated to efficacy of masks and social distancing, airborne transmission, transmission in schools, and COVID-19 in children.
- We found that in the first year of the pandemic, the NIH diverted a small fraction of its budget to COVID-19 research. Future health emergencies will require research funding to pivot in a timely fashion and funding levels to be proportional to the burden of disease in the population.
- The type of research and the clinical/scientific areas studied were based on definitions that may not be collectively exhaustive and mutually exclusive.
- We only reviewed abstracts and did not review the entire funded proposals. There were other barriers to clinical research that were not captured here, including slow institutional review boards and long journal peer-review times.

## Data Availability

All data produced in the present study are available upon reasonable request to the authors

## Patient and Public Involvement

No patient involved in this study.

### Contributors

LB, CD, CMW, and MAM designed the study. LB, CD, AM, SH, CMW, CK, BC, and FM collected the data. LB and CD analysed the data. LB prepared the first draft of the manuscript. LB, CD, AM, SH, CMW, and MAM made critical revisions to the manuscript. All authors reviewed and approved the final draft.

### Competing interests

None declared

### Funding

No Funding was reported for this study.

### Data sharing statement

Data are publicly available.

### Ethics statement

Ethics approval was not sought as the study presents results of an analysis of secondary data and does not involve human participants.

## Acknowledgment

We thank Farah Hashim, Jonathan Teinor, and Karashini Ramamoorthi for their contribution in preparing this manuscript.

## Notes

### Competing Interest Statement

The authors have declared no competing interest.

### Funding Statement

This study did not receive any funding

