## Supplemental tables for "NIH Funding of COVID-19 Research in 2020: a Cross Sectional Study"

**Appendix 1: Definition of Research Types**

| Research Type | Definition | RCDC/NIH Definition |
|---|---|---|
| **Basic Science Research** | Fundamental laboratory or bench research and provides the foundation of knowledge for applied science and encompasses biochemistry, microbiology, physiology, and pharmacology, and their interplay, and involves laboratory studies with cell cultures, animal studies, or physiological experiments[10]<br><br>Adapted from Concept ID: 681833, 1511287 | Concept Name: Basic Science<br>Concept ID: 681833<br>Concept Definition: 0.Research aimed at deriving general knowledge, without a direct application toward solving a specific problem.<br><br>Concept Name: Basic Research Breast Cancer<br>Concept ID: 1511287<br>Concept Definition: 0.Research on the molecular, genetic, biochemical, cellular, structural, immunological, pharmacological mechanisms and factors as they relate to the causation, progression, diagnosis, and treatment of breast cancer. |
| **Clinical Research** | Research conducted with human subjects, or on the material of human origin, in which an investigator directly interacts with human subjects, which are studied to understand health and disease; includes the development of new technologies, mechanism of human diseases, therapy, and clinical trials[11]<br><br>Adapted from Concept ID: 8972 | Concept Name: Clinical Research<br>Concept ID: 8972<br>Concept Definition: research conducted with human subjects or on material of human origin in which an investigator directly interacts with human subjects; includes development of new technologies, mechanism of human diseases, therapy, clinical trials, epidemiology, behavior and health services research. |
| **Translational Research** | Translational research requires interdependence between basic and clinical investigators in both the planning and implementation of research and emphasizes the clinical application of basic research findings with patients and populations.<br>This research has both basic science and clinical components which applies discoveries generated during research in the laboratory and in preclinical studies (basic science research), to the development of trials and studies in humans (clinical research)[12]<br><br>Adapted from Concept ID: 1519620 | Concept Name: Translational Research<br>Concept ID: 1519620<br>Concept Definition: Translational research requires interdependence between basic and clinical investigators in both the planning and implementation of research and emphasizes clinical application of basic research findings with patients and populations. Translational research also applies clinical findings to advance basic research that ultimately may lead to hypothesis-driven clinical trials or prevention and control interventions (from Specialized Programs Of Research Excellence in Prostate Cancer NIH Guide, Volume 23, Number 33, September 16, 1994, RFA: CA-94-031) |

| | | |
|---|---|---|
| **Infrastructure & Education Research** | Infrastructure: Research infrastructure refers to the facilities, resources, and services that are used by the research and innovation community to conduct research and foster innovation in their fields, such as increasing testing capacity, shipping and receiving services, waste management, and utilities, or Vaccine and Treatment Evaluation Units[13]<br><br>Adapted from Concept ID: 1514880 and 1512763<br><br>Education: Research related to training and teaching the general public and/or specific populations focused on improving knowledge of COVID-19 and COVID-19 preventative methods. Using systematic investigation, this research also adopts rigorous and well-defined scientific processes and empirical methods to gather and analyze data in order to solve challenges in education.[14]<br><br>Adapted from Concept ID1514602 | **Education:**<br>Concept Name: Public Health Education<br>Concept ID: 1514602<br>Concept Definition: Health education aimed at the general public<br><br>**Infrastructure:**<br>Concept Name: Research Infrastructure<br>Concept ID: 1514880<br>Concept Definition: 0.Refers to the physical structures needed to conduct research as well as the basic services needed for support, eg shipping and receiving services, waste management, and utilities.<br>Synonym Name: Infrastructure<br><br>Concept Name: Infrastructure Activities<br>Concept ID: 1512763<br>Concept Definition: 0.NIH Emphasis Area: Infrastructure activities are new or expanded programs in the following: Research Training; Shared Instrumentation and Services; Technology Development; Information Technology and Clinical Research. Again, only new or expanded program initiatives should be reported-e.g., an increase in training related only to the increase in stipends should not be reported as an Infrastructure Initiative.<br>Broader Term: Research Infrastructure |
| **Public Health Research** | Public health research tries to improve the health and well-being of people from a population-level perspective including research that addresses mental health and social determinants of health[15]<br><br>Adapted from Concept ID 34019 | Concept Name: Public Health<br>Concept ID: 34019<br>Concept Definition: Branch of medicine concerned with the prevention and control of disease and disability, and the promotion of physical and mental health of the population on the international, national, state, or municipal level. 1.branch of medicine concerned with the prevention and control of disease and disability, and the promotion of physical and mental health of the population on the international, national, state, or municipal level. 2.The science and practice of protecting and improving the health of a community, as by preventive medicine, health education, control of communicable diseases, |

| | | |
|---|---|---|
| | | application of sanitary measures, and monitoring of environmental hazards. (Disability History Museum glossary; http://www.disabilitymuseum.org/glossary.php) Broader Term: Environment and Public Health |
| **Other** | Research that does not fit into the above research types. This includes research that has tangential impacts on COVID-19 knowledge or response but is not primarily focused on COVID-19. | |

**Appendix 2: Definition of Clinical/Scientific Area**

| **Category** | **Definition** | **NIH/RCDC Definition** |
|---|---|---|
| **Cardiology** | Research that analyzes the effects of COVID-19 on the heart, blood vessels, or circulation.<br><br>Adapted from Concept ID: 7226 | Concept Name: Cardiovascular system<br>Concept ID: 7226<br>Concept Definition: 0.The HEART and the BLOOD VESSELS by which BLOOD is pumped and circulated through the body. 1.Relating to the heart and the blood vessels or the circulation. 2.Cardiovascular; pertaining to the heart or blood vessels.<br>Synonym Name: Cardio-vascular |
| **Diagnosis and Testing** | Research involving the development, improvement, and testing of methods and tools for diagnosing, detecting, and monitoring COVID-19 infection.<br><br>Adapted from Concept ID: 11900 | Concept Name: Diagnosis<br>Concept ID: 11900<br>Concept Definition: The determination of the nature of a disease or condition or the distinguishing of one disease or condition from another. Assessment may be made through physical examination, laboratory tests, or the like, and may be assisted by computerized programs designed to enhance the decision-making process. 1.general term for detecting and classifying diseases. 2.The investigation, analysis and recognizing of the presence and nature of disease, condition, or injury from expressed signs and symptoms; also, the scientific determination of any kind; the concise description of characterization of a species where the characteristics of an organism are diagnosed to determine which taxonomic classification is suitable to them. In oncology also: the development, improvement, and testing of methods for cancer detection and staging. |
| **Gastroenterology** | Research that analyzes the effects of COVID-19 infection on the structures and functions of the gastrointestinal tract, including the esophagus, stomach, small intestine, and large intestine (colon, rectum, and anus) and associated digestive organs (liver, gallbladder, and pancreas) | Concept Name: Gastroenterology<br>Concept ID: 17163<br>Concept Definition: 0.A subspecialty of internal medicine concerned with the study of the physiology and diseases of the digestive system and related structures (esophagus, liver, gallbladder, and pancreas). |

| | | |
|---|---|---|
| | Adapted from Concept ID: 17163, 17178 | Concept Name: Gastrointestinal Diseases<br>Concept ID: 17178<br>Concept Definition: 0.Diseases in any segment of the GASTROINTESTINAL TRACT from ESOPHAGUS to RECTUM. 1.disorder in any segment of the gastrointestinal tract from the esophagus to the rectum. 2.RAEB: Diseases of the digestive tract (oral cavity to anus) and associated organs (salivary glands, liver, pancreas). 3.RAEB: Use for studies in which the focus is on the digestive tract (oral cavity to anus) and associated organs (salivary glands, liver, pancreas). For most digestive organs there will be no problem; however, liver is frequently used in studies of carcinogens for which it is not normally the target organ. In the latter type case do not code SIC 36. (NCI) |
| **Geriatrics** | Research concerned with the physiological and pathological aspects of the aged, including the clinical problems of senescence and senility. This includes clinical research involving human subjects above the age of 65 years.<br><br>Adapted from Concept ID: 17469 | Concept Name: Geriatrics<br>Concept ID: 17469<br>Concept Definition: 0.The branch of medicine concerned with the physiological and pathological aspects of the aged, including the clinical problems of senescence and senility. 1.field of medicine concerning elderly human health. 2.The branch of medical science that deals with diseases and problems specific to elderly people. |
| **Immunology** | Research of the immune system and its reaction to, as well as its malfunctions in response to COVID-19 infection. This includes research that pertains to the identification and characterization of immune factors; immune physiology; diseases of the immune system in conjunction with COVID-19 infection or complications.<br><br>Adapted from Concept ID: 152036 | Concept Name: Immunology<br>Concept ID: 152036<br>Concept Definition: The occupation or discipline. 1.Immunology is the study of the immune system and its reaction to pathogens, as well as its malfunctions (autoimmune diseases, allergies, rejection of organ transplants). (from Wikipedia) 2.RAEB: Use for any aspect of immunology: identification and characterization of immune factors; immune system development; immune physiology; immunotherapy; immunodiagnosis; tumor or virus antigen studies; vaccine research; diseases of the immune system (immunodeficiencies, autoimmunity, hematopoietic system neoplasia). Not used for research tools such as antibody tagging if the study is otherwise unrelated to immunology. (NCI)<br>Broader Term: Biological Sciences |

| | | |
|---|---|---|
| **Impacts on Other Diseases** | Research analyzing the impact of the COVID-19 infection and pandemic control measures on the pre-existing disease or condition the patient is diagnosed with.<br><br>Adapted from Concept ID: 9599 | Concept Name: Complication<br>Concept ID: 9566<br>Concept Definition: 0.something that introduces usually unexpected difficulties, problems, or changes. 1.Any disease or disorder that occurs during the course of (or because of) another disease. |
| **Maternal Health** | Research that analyzes the impact of COVID-19 infection, complications, or pandemic control measures on maternal health including pregnancy, prenatal care, labor and delivery, and childcare.<br><br>Adapted from Concept ID: 1513012, and 33052 | Concept Name: Maternal and Child Health<br>Concept ID: 1513012<br>Usually involves maternal factors (and efforts to modify these factors) that may affect the health of the child or fetus: smoking or exposure to drugs or toxic chemicals during pregnancy, maternal/fetal immunologic interactions. Also use for genetic counseling, pregnancy and/or nursing effects on maternal health.<br><br>Concept Name: Prenatal Care<br>Concept ID: 33052<br>Care provided the pregnant woman in order to prevent complications, and decrease the incidence of maternal and perinatal mortality. |
| **Non-Pharmaceutical Interventions** | Research regarding investigating the implementation and/or efficacy of non-pharmacological measures to address the COVID-19 pandemic, including programs designed to prevent and control the spread of infection.<br><br>Adapted from Concept ID: 85557 | Concept Name: Infection Control<br>Concept ID: 85557<br>Concept Definition: Programs of disease surveillance, generally within health care facilities, designed to investigate, prevent, and control the spread of infections and their causative microorganisms. |
| **Nephrology** | Research that analyzes the effects of COVID-19 infection on the structures, functions, and diseases of the renal system.<br>Adapted from Concept ID: 27712 | Concept Name: Nephrology<br>Concept ID: 27712<br>Concept Definition: 0.A subspecialty of internal medicine concerned with the anatomy, physiology, and pathology of the kidney. |
| **Neurology** | Research that analyzes the effects of COVID-19 infection on the structures, functions, and diseases of the nervous system including effects on senses. | Concept Name: Neurology<br>Concept ID: 27855<br>Concept Definition: 0.A medical specialty concerned with the study of the structures, functions, and diseases of the nervous system. 1.the branch of |

| | | |
|---|---|---|
| | Adapted from Concept ID: 27855, 36658 | medical science that deals with the study of structure, function, and diseases of the nervous system;<br><br>Concept Name: Esthesia<br>Concept ID: 36658<br>Concept Definition: 0.Transduction of physical or chemical changes in the external or internal environment into nerve impulses by specialized receptors, transmission of these impulses by afferent neurons to the effectors, either directly or through the CNS. 1.transduction of stimuli from outside the body and those within the body into nerve impulses by receptors, and the transmission of these impulses by afferent neurons to the cerebral cortex.<br>Synonym Name: Sensation |
| **Other Research** | Grants that do not fall into other clinical/scientific areas. | |
| **Pediatric Health** | Research concerned with maintaining health or providing medical care to children from birth to adolescence in the context of COVID-19 infection, complications, or control measures.<br><br>Adapted from Concept ID 30755, 1578 | Concept Name: Pediatrics<br>Concept ID: 30755<br>Concept Definition: 0.A medical specialty concerned with maintaining health and providing medical care to children from birth to adolescence.<br><br>Concept Name: Adolescence<br>Concept ID: 1578<br>Concept Definition: 0.period of life beginning with the appearance of secondary sex characteristics and terminating with the cessation of somatic growth; typically between 12 and 20 years of age; when school grades are referenced, this age group is typically grade 5 or 6 and above; also index with appropriate human and clinical research terms. 1.Adolescence is the time period between the beginning of puberty and adulthood. 2.The period of life beginning with the appearance of secondary sex characteristics and terminating with the cessation of somatic growth. The years usually referred to as adolescence lie between 13 and 18 years of age. |
| **Pharmaceutical Interventions** | These are preclinical and clinical studies analyzing the nature, properties, and actions of drugs as therapeutics for COVID-19 infection. This includes research involving the creation and | Concept Name: New Agents<br>Concept ID: 1518316<br>Concept Definition: 0.Research into new physical or chemical means to treat disease.<br>Broader Term: Funding Category |

| | | |
|---|---|---|
| | testing of new therapeutics, interventions, vaccines, and repurposing of prior FDA approved therapeutics to treat infection, alleviate symptoms, or offer prophylaxis against COVID-19.<br><br>Adapted from Concept ID: 1518316, 31330 | Concept Name: Pharmacology<br>Concept ID: 31330<br>Concept Definition: 0.The study of the origin, nature, properties, and actions of drugs and their effects on living organisms. 1.the biological effects of drugs in living organisms or tissues; use this term mainly for intended, desired effects; for harmful or undesired effects, see DRUG ADVERSE EFFECT or TOXICOLOGY. 2.Pharmacology is the study of drugs in all their aspects. It is concerned with the art and science of the preparation, compounding, and dispensing of drugs. (Pharmacology Glossary; http://www.bumc.bu.edu)<br>Broader Term: Biological Sciences |
| **Pulmonology** | Research that analyzes the effects of COVID-19 infection on the respiratory system and respiration disorders.<br><br>Adapted from Concept ID 35204 | Concept Name: Respiration Disorders<br>Concept ID: 35204<br>Concept Definition: Diseases of the respiratory system in general or unspecified or for a specific respiratory disease not available. |
| **Risk Factor Analysis** | Research that provides qualitative or quantitative estimation of susceptibility to COVID-19 infection and/or adverse outcomes based on the presence of risk factors, herein defined as inherited, environmental, or behavioral characteristics that affect COVID-19 infection, symptoms, and outcome.<br><br>Adapted from Concept ID 86930, 35648, 12655 | Concept Name: Risk<br>Concept ID: 35647<br>Concept Definition: 0.The probability that an event will occur. It encompasses a variety of measures of the probability of a generally unfavorable outcome. 1.Risk is the potential future harm that may arise from some present action. It is often combined or confused with the probability of an event which is seen as undesirable. (from WIkipedia)<br><br>Concept Name: Risk Assessment<br>Concept ID: 86930<br>Concept Definition: 0.The qualitative or quantitative estimation of the likelihood of adverse effects that may result from exposure to specified health hazards or from the absence of beneficial influences. (Last, Dictionary of Epidemiology, 1988) 1.The qualitative or quantitative estimation of the likelihood of adverse effects that may result from exposure to specified health hazards or from the absence of beneficial influences. |

| | | |
|---|---|---|
| | | Concept Name: Risk Factors<br>Concept ID: 35648<br>Concept Definition: 0.An aspect of personal behavior or lifestyle, environmental exposure, or inborn or inherited characteristic, which, on the basis of epidemiologic evidence, is known to be associated with a health-related condition considered important to prevent.<br><br>Concept Name: Disease susceptibility<br>Concept ID: 12655<br>Concept Definition: 0.A constitution or condition of the body which makes the tissues react in special ways to certain extrinsic stimuli and thus tends to make the individual more than usually susceptible to certain diseases. 1.factors that affect the probability or predisposition of an individual to the development of a disease(s) or disorder(s). |
| **Social Determinants of Health** | Social determinants of health (SDOH) are the conditions in the environments where people are born, live, learn, work, play, worship, and age that affect a wide range of health, functioning, and quality-of-life outcomes and risks. Research in this area identifies the effect of social determinants on people's health in the context of COVID-19 infection and preventive methods[16]<br><br>Adapted from Concept ID 1171307, 37470, 26192 | Concept Name: health disparity<br>Concept ID: 1171307<br>Concept Definition: 0.a population-specific difference in the presence of disease, health outcomes or access to care.<br><br>Concept Name: Infectious Disease Epidemiology<br>Concept ID: 1512717<br>Concept Definition: 0.Epidemiology as it relates to infectious diseases.<br><br>Concept Name: Medical Sociology<br>Concept ID: 37470<br>Concept Definition: 0.The study of the social determinants and social effects of health and disease, and of the social structure of medical institutions or professions. 1.Medical sociology is the study of individual and group behaviors with respect to health and illness. Medical sociology is concerned with individual and group responses aimed at assessing |

| | | |
|---|---|---|
| | | well-being, maintaining health, acting upon real or perceived illness, interacting with health care systems, and maximizing health in the face of physiologic or functional derangement. It also analyzes the impact of the psychological conditions resulting from our environment on our health.<br><br>Concept Name: Minority Groups<br>Concept ID: 26192<br>Concept Definition: 0.A subgroup having special characteristics within a larger group, often bound together by special ties which distinguish it from the larger group. 1.A minority is a group that is outnumbered by persons who do not belong to it, often people with different nationality, religion, culture or lifestyle from that of the mainstream in the society. (Wikipedia) 2.RAEB: Racial or ethnic groups officially recognized by the U.S. government as minority populations. |
| **Transmission** | Research that describes or models the transmission of COVID-19 focusing on where and how transmission occurs. This includes research specifically addressing when infected individuals are most contagious, disease duration, and the period in which infectivity or illness resolves.<br><br>Adapted from Concept ID 242781, 242649 | Concept Name: Disease transmission<br>Concept ID: 242781<br>Concept Definition: 0.The transmission of infectious disease or pathogens. When transmission is within the same species, the mode can be horizontal (DISEASE TRANSMISSION, HORIZONTAL) or vertical (DISEASE TRANSMISSION, VERTICAL). 1.transmission of an infectious disease by direct contact with an affected individual, the individual's discharges or by indirect means such as by a vector. 2.The transmission of infectious disease or pathogens. When transmission is within the same species, the mode can be horizontal (disease transmission, horizontal) or vertical (disease transmission, vertical). (NCI)<br><br>Concept Name: Horizontal Disease Transmission<br>Concept ID: 242649<br>Concept Definition: 0.The transmission of infectious disease or pathogens from one individual to another in the same generation. |

| | | |
|---|---|---|
| **Virology** | Research analyzing the characteristics of the virus SARS-CoV-2, including molecular viral components pertaining to replication, infectivity, and genetic variability.<br><br>Adapted from Concept ID 597650, 42774, 34848, 597652, 597653 | Concept Name: virus characteristic<br>Concept ID: 597650<br>Concept Definition: features that help to identify, distinguish or describe recognizably; classification systems of viruses; includes infection routes, staining patterns, replication requirements, etc.<br><br>Concept Name: Virus Replication<br>Concept ID: 42774<br>Concept Definition: 0.The process of intracellular viral multiplication, consisting of the synthesis of PROTEINS; NUCLEIC ACIDS; and sometimes LIPIDS, and their assembly into a new infectious particle. 1.process of forming progeny virus from input virus; involves expression and replication of viral genomic nucleic acid and the assembly of progeny virus particles.<br><br>Concept Name: Virus Receptors<br>Concept ID: 34848<br>Concept Definition: 0.Specific molecular components of the cell capable of recognizing and interacting with a virus, and which, after binding it, are capable of generating some signal that initiates the chain of events leading to the biological response. 1.viruses must first bind to their target cell's surface before infection can proceed; this is mediated by specific molecular receptors for certain viral antigens, many of which have other known functions; e.g., the MHC-II receptor CD4 is also an HIV receptor. 2.Cell surface molecules that are capable of interacting with virus particles, thereby mediating their entry into the cell or otherwise eliciting a cellular response.<br><br>Concept Name: virus infection mechanism<br>Concept ID: 597653<br>Concept Definition: 0.multi-step process by which a virus binds to, enters, and replicates within a host |

| | | |
|---|---|---|
| | | cell; includes both surface and intracellular interactions between host and virus.<br><br>Concept Name: virus genetics<br>Concept ID: 597652<br>Concept Definition: 0.heredity, especially the mechanisms of hereditary transmission and the variation of inherited characteristics among a virus or viruses; the genetic constitution of viruses. 1.The branch of science concerned with the means and consequences of viral transmission and generation of the components of biological inheritance. |
